# Parents’ and guardians’ views on the acceptability of a future Covid-19 vaccine: a multi-methods study in England

**DOI:** 10.1101/2020.09.16.20188227

**Authors:** Sadie Bell, Richard Clarke, Sandra Mounier-Jack, Jemma L Walker, Pauline Paterson

## Abstract

**Background:** The availability of a COVID-19 vaccine has been heralded as key to controlling the COVID-19 pandemic. COVID-19 vaccination programme success will rely on public willingness to be vaccinated.

**Methods:** We used a multi-methods approach – involving an online cross-sectional survey and semi-structured interviews - to investigate the views of parents’ and guardians’ on the acceptability of a future COVID-19 vaccine. 1252 parents and guardians (aged 16+ years) who reported living in England with a child aged 18 months or under completed the survey. Nineteen survey respondents were interviewed.

**Findings:** Most participants reported they would definitely accept or were unsure but leaning towards accepting a COVID-19 vaccine for themselves (*Definitely 55.8%; Unsure but leaning towards yes 34.3%)* and their child/children *(Definitely 48.2%; Unsure but leaning towards yes 40.9%)*. Participants were more likely to accept a COVID-19 vaccine for themselves than for their child/children. Less than 4% of participants reported that they would definitely not accept a COVID-19 vaccine for themselves or their children. Participants that self-reported as Black, Asian, Chinese, Mixed or Other ethnicity were almost 3 times more likely to reject a COVID-19 vaccine for themselves and their children than White British, White Irish and White Other participants. Respondents from lower income households were also more likely to reject a COVID-19 vaccine.

The main reason for vaccine acceptance was for self-protection from COVID-19. Common concerns were around COVID-19 vaccine safety and effectiveness, which were largely prompted by the newness and rapid development of the vaccine.

**Conclusion:** To alleviate concerns, information on how COVID-19 vaccines are developed and tested, including their safety and efficacy, must be communicated clearly to the public. To prevent Inequalities in uptake, it is crucial to understand and address factors that may affect COVID-19 vaccine acceptability in ethnic minority lower-income groups who are disproportionately affected by COVID-19.

## Background

Over 20.6 million cases of coronavirus (COVID-19) and almost 750,000 deaths have been reported globally since the start of the COVID-19 pandemic to 13^th^ August 2020 [1]. In addition to the global mortality and morbidity costs attributed to COVID-19, the pandemic and restrictions implemented to slow COVID-19 spread (i.e. lockdown, quarantine, curfew and social distancing measures) have resulted in devastating social and economic consequences. The pandemic has exacerbated global inequalities, with the world bank predicting that 71 million people will be pushed into extreme poverty in 2020 [2],

According to COVID-19 mortality figures, the UK is one of the worst affected countries in the world, with over 310,000 cases and 41,000 deaths reported as of 13^th^ August 2020 [1, 3]. In efforts to slow the spread of COVID-19 in the UK, stringent restrictions on movement were enforced on 23^rd^ March 2020, with the general public directed to only leave their homes to: shop for basic essentials, take one form of exercise per day, access medical care or help a vulnerable person, and to travel to and from work if absolutely necessary (where unable to work from home) [4], People most at-risk from COVID-19 were asked to protect themselves by shielding from 23^rd^ March 2020: staying at home at all times, for at least 12 weeks. In addition, schools were closed to all but children of key workers or vulnerable children [4],

Although restrictions to slow COVID-19 spread have eased in many countries, including the UK, day-to-day life globally remains distinctly marked by the COVID-19 pandemic, and measures to prevent COVID-19 spread [5]. A COVID-19 vaccine has been heralded as key to ending the pandemic [6]. With an accelerated process, in which development stages take place in parallel, scientists aim to produce a COVID-19 vaccine within 12-18 months, at an anticipated cost of at least $3 billion for vaccine development, manufacture and delivery [7, 8]. In the UK, a Vaccine taskforce has been created to ‘drive forward, expedite and co-ordinate efforts to research and then produce a coronavirus vaccine and make sure one is made available to the public as quickly as possible’ [6]. On 20^th^ July 2020, the UK government secured early access to 90 million doses of promising COVID-19 vaccines, ‘giving the UK the most likely chance of getting access to a safe and effective vaccine at the quickest speed’ [9].

In planning for the near-future availability of a COVID-19 vaccine, as well as focusing on how to deliver COVID-19 vaccine programmes and ensure equitable vaccine allocation globally [10], it is crucial to explore the acceptability of a COVID-19 vaccine to the public. The success of any COVID-19 vaccination programme will depend on public willingness to receive the vaccination.

The priority groups to receive a future COVID-19 vaccine in the UK are frontline National Health Service (NHS) workers and social care workers, and those at increased risk of COVID-19 (i.e. due to age or other risk factors, including underlying health conditions). It is not currently known when or if children or adults not in risk groups for COVID-19 will be eligible for a COVID-19 vaccine. Vaccinating young and healthy members of the population may help to prevent COVID-19 transmission to high-risk groups. We conducted a multi-methods study to investigate the views of parents and guardians in England on the acceptability of a future COVID-19 vaccine, looking at their views on receiving a COVID-19 vaccine for themselves and their children.

## Methods

We used a multi-methods approach - combining qualitative and quantitative methods - with the aim of gaining a more complete insight into the acceptability of a future COVID-19 vaccine. The study utilised a cross-sectional questionnaire survey and semi-structured interviews, to quantify the prevalence of different views on COVID-19 vaccine acceptability, and to explore reasons behind these views. The questions asked regarding COVID-19 vaccine acceptance were part of a larger study exploring parents’ and guardians’ views and perceptions towards childhood vaccinations and use of NHS general practice services for childhood vaccination during the COVID-19 pandemic

### Cross-sectional survey

#### Recruitment

Survey recruitment took place between 19th April and 11th May 2020. Eligible participants were required to be (1) a parent or guardian of a child (or children) aged 18 months or under, (2) a resident of England, and (3) aged 16+ years.

We utilised an online social media strategy to recruit survey participants. This strategy involved the creation of a Facebook page dedicated to the study with a single post advertising the study, outlining the eligibility criteria for participation, and including the hyperlink to the survey. An email was then sent to organisers of 284 baby and toddler play groups in England. This contact email included a study summary and a request to share the advertising post through the groups various means of communication (e.g. Facebook, WhatsApp, Twitter or an email list). In addition to this, Facebook’s paid promotion feature was used to target the survey at eligible potential participants.

#### Measures

The survey included demographic questions concerning age, gender, household income, employment, marital status of participants, and number and age of children. Two questions were designed to capture acceptance of a new COVID-19 vaccination. The text of the questions read *“If a new coronavirus (COVID-19) vaccine became available would you accept the vaccine for yourself?”* and *“If a new coronavirus (COVID-19) vaccine became available would you accept the vaccine for your child/children?’*. A 4-point Likert scale was used to encourage participants to take a stance on the vaccine (rather than selecting a ‘do not know’ answer). The Likert scale options were *“Yes, definitely”, “Unsure but leaning towards yes”, “Unsure but leaning towards no”* and *“No, definitely not”*. Open-text boxes were included for participants to explain their responses to the two questions on vaccine acceptability for themselves and for their children. Participants could give more than one reason in the open-text boxes.

#### Analysis

A paired samples t-test was used to compare acceptance of a COVID-19 vaccine for self and for the participant’s child. Two subsequent logistic regressions were then conducted to determine the demographic factors associated with rejection of the COVID-19 vaccine for both self-vaccination and that of the participant’s child. Alpha was set to 0.05 for all tests. Age, household income, ethnicity, location, and employment were included as predictive variables in the logistic regression models. In the logistic regression model for child vaccination, number of children was also included as a predictive variable.

To perform the logistic regressions ethnicity was dichotomised into *‘White’ (*i.e. White British, White Irish and White Other participants) and *‘Black, Asian or minority ethnic (BAME)’* (i.e. Black, Asian, Chinese, Mixed or Other ethnicity). Household income brackets were reduced to a *“low income”* (<£35,000), *“medium income”* (£35,000 - £84,999) and *“high income”* (>£85,000), and the vaccine acceptance variables were dichotomised into *“likely to accept”* (those that answered *Yes, definitely* or *Unsure but leaning towards yes)* and *“likely to reject”* (those that answered *No, definitely* not or *Unsure but leaning towards no)*.

Free text responses were analysed thematically in Microsoft Excel. Coding schemes were produced based on the content of the free-text comments.

### Semi-structured interviews

On completing the survey, participants were asked to provide their contact details if interested in taking part in a follow-up semi-structured interview. Participants that had expressed interest were purposively selected based on a range of characteristics, including ethnicity, household income, and geographical location.

Participants were emailed an information sheet, fully detailing the study objectives and explaining all aspects of participation, including the right to withdraw from the research. Written informed consent was obtained from each participant. Interviews lasted approximately 30 minutes and were conducted over the phone. Topic guides, shaped around the content of the questionnaire, were used to assist the interviews. Interview participants received a £10 gift voucher as a thank you for their time and contribution. The interviews took place between 27^th^ April and 27^th^ May 2020.

Interviews were transcribed verbatim and analysed thematically in NVivo12 using the stages outlined by Braun and Clarke [11]: data familiarisation, coding and theme identification and refinement. To enhance the rigour of the analysis, coding approaches and data interpretations were discussed between SB, RC, PP and SM-J.

### Public involvement

We gained feedback from parents with young children, including children aged 18 months or under, in the refinement of the survey questions and layout. This involvement aimed to increase the user-friendliness and appropriateness of the survey.

## Findings

1252 parents and guardians completed the survey (see participant characteristics in Table 1). Most survey participants were female (95.0%), raising a child/children with a partner (97.0%), and identified as being White British, White Irish or White Other (94.1%). The age range of participants was 18-48 years (Mean=32.95, SD=4.565). Median household income was reported as £55,000-£64,999. Of the survey participants, 1049 (83.8%) left free-text responses to the open text survey questions.

**Table 1.**
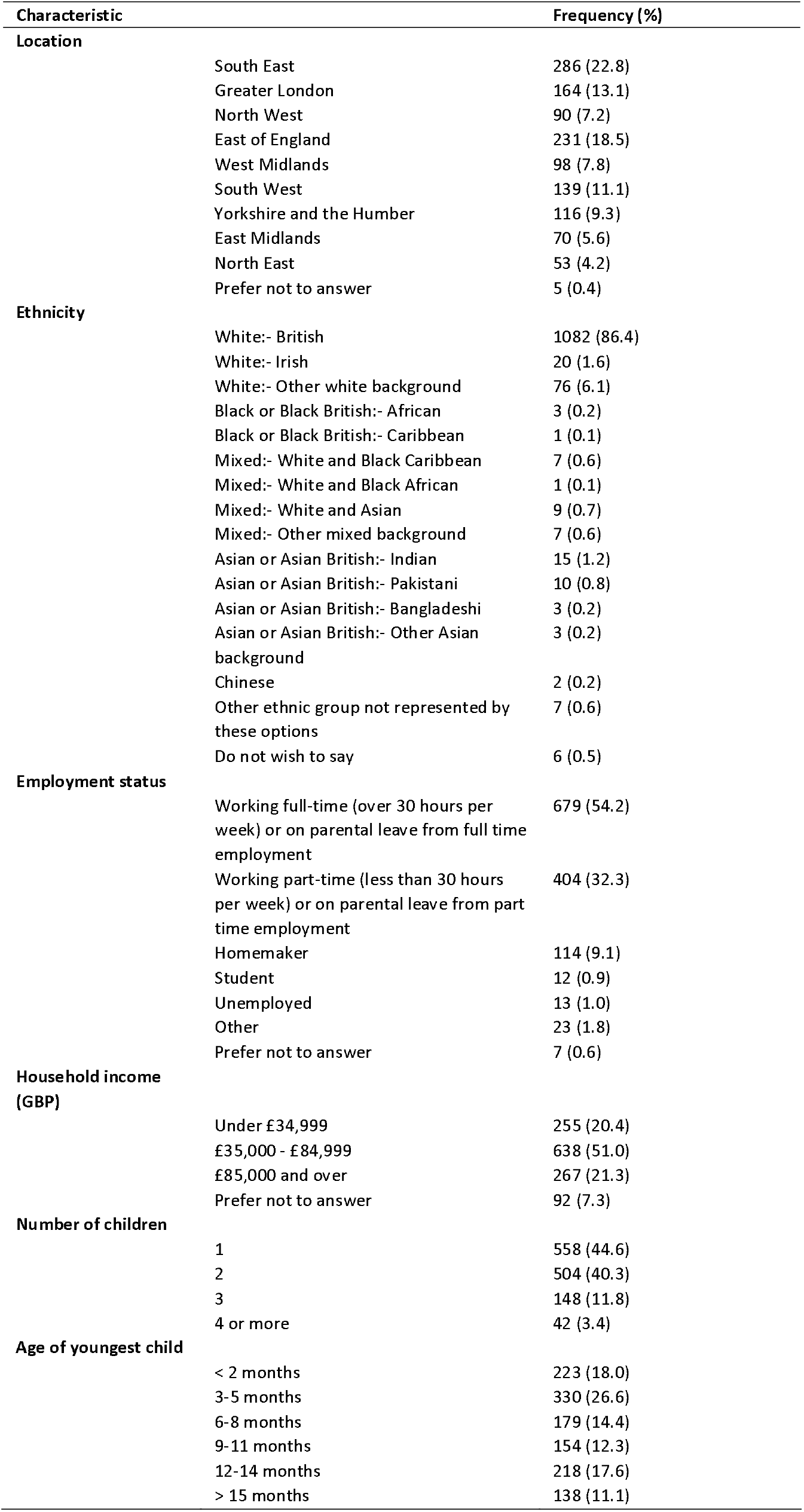

43.3% of survey participants (n=530) provided their details to be contacted for a follow-on interview. In total, 61 parents were contacted to participate. Of these 19 took part in interviews (18 women and one man), 39 did not respond to recruitment emails, two responded initially but did not follow through with an interview. The characteristics of interviewees are outlined in Table 2.

**Table 2.**
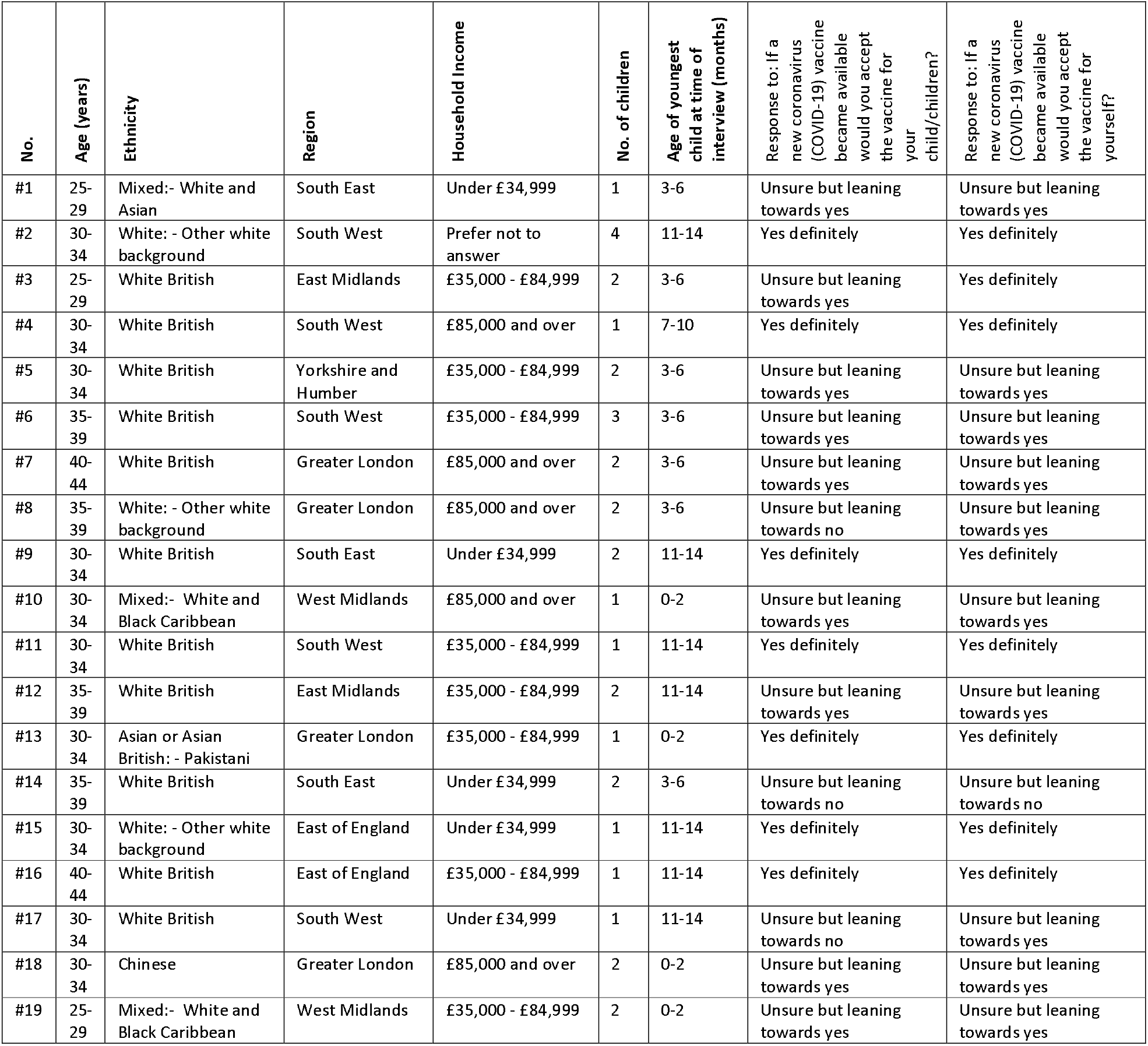

*Quantitative findings - Acceptability of a future COVID-19 vaccination* A high proportion of survey participants reported that they would definitely accept or were unsure but leaning towards accepting a COVID-19 vaccine for themselves (*Yes, definitely* 55.8%, n=699; *Unsure but leaning towards yes* 34.3%, n=429) and their child/children (*Yes, definitely* 48.2%, n=604; *Unsure but leaning towards yes* 40.9%, n=512) (Table 3). Less than 1 in 10 participants reported that they were unsure but leaning towards rejecting or would definitely reject a COVID-19 vaccine for themselves (*Unsure but leaning towards no 6.2%, n=78; No, definitely not 3.7%, n=46)* and their child/children (*Unsure but leaning towards no 7.4%, n=93; No, definitely not 3.4%, n=43)*.

Participants were more likely to say that they would definitely accept or were leaning towards accepting a COVID-19 vaccine for themselves (mean = 2.42, SD =.768) than for their child/children (mean = 2.34, SD =.761), *t (1251) = 7.636 p<.001*.

**Table 3.**
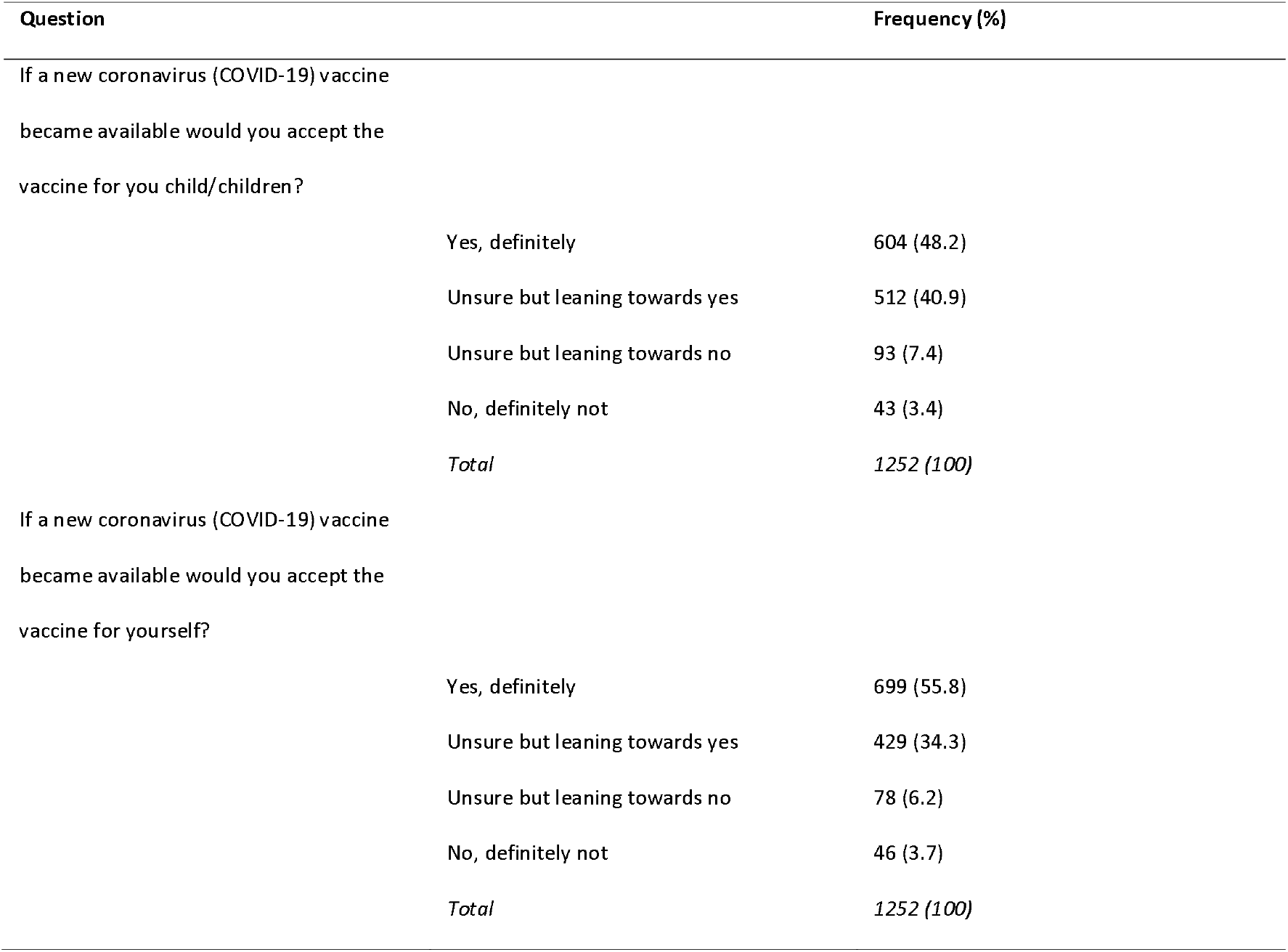

| Question | Frequency (%) |
| --- | --- |
| If a new coronavirus (COVID-19) vaccine became available would you accept the vaccine for you child/children? |  |
| Yes, definitely | 604 (48.2) |
| Unsure but leaning towards yes | 512 (40.9) |
| Unsure but leaning towards no | 93 (7.4) |
| No, definitely not | 43 (3.4) |
| *Total* | *1252 (100)* |
| If a new coronavirus (COVID-19) vaccine became available would you accept the vaccine for yourself? |  |
| Yes, definitely | 699 (55.8) |
| Unsure but leaning towards yes | 429 (34.3) |
| Unsure but leaning towards no | 78 (6.2) |
| No, definitely not | 46 (3.7) |
| *Total* | *1252 (100)* |

*Survey open-text and interview findings: main reasons for accepting the vaccine (Figure 1)*

Of the survey participants that stated they would definitely accept or were unsure but leaning towards accepting a COVID-19 vaccine for themselves (n=1128), 897 left an open-text reason (79.5%). 231 participants did not complete the open text (20.5%). Of the participants that stated they would definitely accept or were leaning towards accepting a COVID-19 vaccine for their child (n=1116), 925 (82.9%) gave a reason for definitely accepting or leaning towards accepting a COVID-19 vaccine for their child. 191 participants did not complete the open text (17.1%).

All interviewees said that they would definitely accept or were unsure but leaning towards accepting COVID-19 vaccine for themselves in their survey responses. Sixteen interviewees said they would definitely accept or were leaning towards accepting a COVID-19 vaccine for their children in their survey responses (see Table 2). Interestingly, one interviewee (#19) that was leaning towards accepting the vaccinate at the time of completing the survey was leaning towards refusing the vaccination (for herself and her child) at the time of interview.

The following reasons were given for COVID-19 vaccine acceptance for self and for child/children, in order of how often they were mentioned by survey participants and importance to interviewees.

### To protect self and others

Of survey participants expressing positive intentions to vaccinate and leaving an open-text response, the most prevalent reason was to provide protection from COVID-19 to the person being vaccinated (*for self* n=380, 42.4*%; for child* n=393, 42.5%), followed by protecting other people (*for self* n=213, 23.7%; *for child* n=180, 19.5%), including family members *(for self* n =109, 12.2%; *for child* n=49, 5.3%). Participants also reported that they would vaccinate to protect someone known to them in a risk group for COVID-19 (*for self* n=74, 8.2%; *for child* n=29, 3.1%). 5.0% (n=45) of survey participants specifically mentioned that they wanted to receive the vaccine to stay healthy to look after their child/children. Interviewees also corroborated these findings, highlighting that vaccinating would make them feel safer in visiting older family members.

> *‘….for us, I definitely want that. I’d want us protected as a family. You know, my parents, his [partner’s] parents are ageing, so we’d be going to see them, and I don’t want to put them at risk. So, certainly, I think it’s really important.’ (Interview #13)*

Several survey participants cited that they would vaccinate as they were key workers (e.g. involved in health and social care, education and childcare, or the food sector) and in frequent contact with other people (n=60, 6.7%). Health and social care workers particularly noted a need to vaccinate to protect themselves and ‘at risk’ patients and clients.

### Vaccination as a route to ‘returning to normal life’

For several interviewees, the availability of a vaccine was viewed as the only way of ending social-distancing measures and returning to normal life. Interviewees talked about lockdown as being financially unsustainable, and detrimental to physical and mental wellbeing and children’s educational and social development. Interviewees that were shielding during the pandemic were particularly keen to accept a future vaccine, with one parent who had looked into entering a COVID-19 vaccine trial saying *they ‘would be first at the door’*.

Survey participants also indicated that they would be vaccinated (n=33, 3.7%) and get their children vaccinated (n=24, 2.6%) as a means of stopping the need for social distancing.

### Trust and belief in the importance of vaccination

Survey participants indicated that they would receive the vaccine for themselves (n=46, 5.1%) or their child/children (n=65, 7.0%) if it was recommended by the government or healthcare workers. Survey participants cited their belief in the importance of vaccination as a reason to receive the vaccination themselves (n=35, 3.9%) or to vaccinate their children (n=50, 5.4%).

A proportion of survey participants specifically reported trust in vaccinations, science or healthcare workers as a prompt to vaccinate their child/children (n=38, 4.1%) and themselves (n=24, 2.7%).

### Less risk in vaccinating adults

Interviewees discussed a greater willingness to ‘risk’ receiving the vaccination for themselves than for their children due to safety concerns. Survey participants also reported the perception that it was less risky to vaccinate adults than children (n=29, 3.2%) as the vaccine trials were conducted in adults.

**Figure 1.**
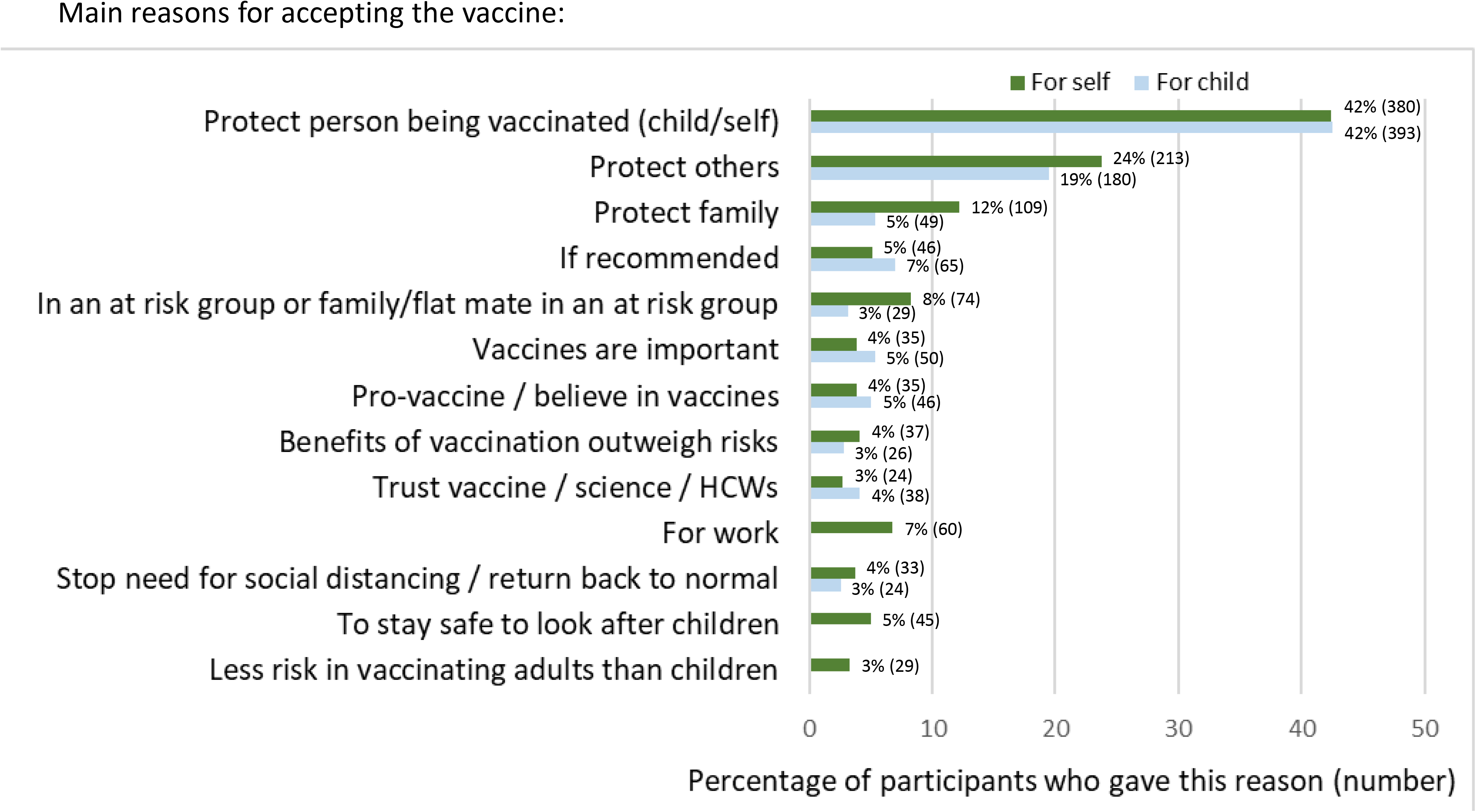

Survey open-text and interview findings: main reasons for <u>not</u> accepting the vaccine (Figure 2)

Of the participants that stated they would definitely reject or were unsure but leaning towards rejecting a COVID-19 vaccine for themselves, 100 left an open-text reason (80.6%) and 24 did not give an open-text reason (19.4%). 124 (91.2%) gave a reason for definitely rejecting or leaning towards rejecting a COVID-19 vaccine for their child and 12 did not give an open-text reason (8.8%). Three interviewees reported that they were unsure but leaning towards rejecting a COVID-19 vaccine for their child/children (see Table 2).

The following reasons were given for COVID-19 vaccine rejection by survey participants and interviewees.

### Vaccine safety and effectiveness concerns

The most common concerns expressed by survey participants about a COVID-19 vaccine were around vaccine safety (*for self* n=49, 49.0%; *for child* n=77, 62.1%) and effectiveness (*for self* n=11, 11.0%; *for child* n=10, 8.1%), which were largely prompted by the newness and rapid development of the vaccine (*for self* n=50, 50.0%; *for child* n=84, 67.7%), and the newness of COVID-19 (*for self* n=3, 3.0%; *for child* n=5, 4.0%). Participants were worried that the development process might be too rapid, not allowing enough time for adequate testing to confirm the vaccine’s short and long-term safety, and also vaccine effectiveness.

These concerns were echoed by interviewees who had reservations around how a vaccine developed in a fraction of the usual timeline for vaccine development could be confirmed as

> *‘I’m very pro-vaccines in general, but my main concern was as long as it hasn’t been rushed through. As long as we know it’safe and all the proper tests have been done, and, you know, when you sort of heard things in the news like well, it’s been 18 years and they haven’t developed a SARS vaccines, or whatever it was that was sort of banded around yesterday, it sort of makes you go, why is that? So, I think that if a vaccine was to come out like this one could do in a hurry it need to have associated information to how this has been able to happen so quickly.’ (Interviewee #19)*

Some survey participants and interviewees wanted to ‘wait-and-see’ if the vaccination was safe before being vaccinated themselves, or getting their children vaccinated. One interviewee discussed preferring for her family to remain in lockdown for longer (i.e. living for a longer duration with the restrictions implemented on 23^rd^ March 2020), to allow more time for vaccine development and safety testing. This interviewee acknowledged this situation as not a financially viable option for those unable to work from home.

### Children less at risk of COVID-19

Some survey participants expressed a lack of benefit to vaccinating their children, citing that children are hardly affected, are at lower risk of severe COVID-19 infection than adults (n=24,19.4%), and less likely to catch or transmit COVID-19 (n=4, 3.2%). Participants and interviewees discussed the decision to vaccinate their children in terms of balancing potential advantages and disadvantages. The perception that children are less at risk of COVID-19 was combined with safety concerns as a reason not to vaccinate.

> *‘Any vaccine for covid-19 will have been produced in a huge hurry, possibly bypassing some of the normal processes and procedures that ensure safety. As small children have a very low risk of being seriously ill from the virus, I’m not sure I would risk it. I would encourage my mum, who is 70 with underlying health conditions, to take it though as the risk/reward is different.’ (Survey participant #1095)*

Several survey participants indicated that they would be more likely to get their child/children vaccinated if this was beneficial in reducing COVID-19 spread to older people and clinical risk groups.

### Low perception of risk

Some survey participants indicated not needing the vaccine as they (n=16,16.0%) and their child/children (n=4, 3.2%) were ‘fit and healthy’ and not in an at-risk group. 2% of participants (n=2) indicated that it was better to prioritise ‘at-risk’ groups for vaccination.

### Already had COVID-19

A small number of survey participants indicated that they would not accept a COVID-19 vaccine for themselves or their child/children as they (n=6, 6.0%) or their child (n=4, 3.2%) had already had COVID-19.

### Need for transparency to make an informed vaccine decision

To be able to make an informed choice about vaccination, parents expressed a need for transparent information on vaccine development, vaccine efficacy and vaccine safety. A lack of vaccine information at present was given as a reason not to accept the vaccine for a minority of survey participants for their children (n=6, 4.8%) and for themselves (n=1, 1.0%). One interviewee also discussed that the information should come from trusted sources such as healthcare professionals, the NHS and Public Health England.

*‘…if the NHS is telling you something you’re more likely to believe it, whereas if it’s a politician, you know, you’ve got much more incentive to think, “Well, are they telling the truth?” (Interviewee #17)*

### Lack of trust

A small number of survey participants stated a lack of trust in vaccinations, science or the medical profession as a reason to not accept a vaccine *(for self* n =4, 4.0*%; for child* n =2, 1.6%).

**Figure 2.**
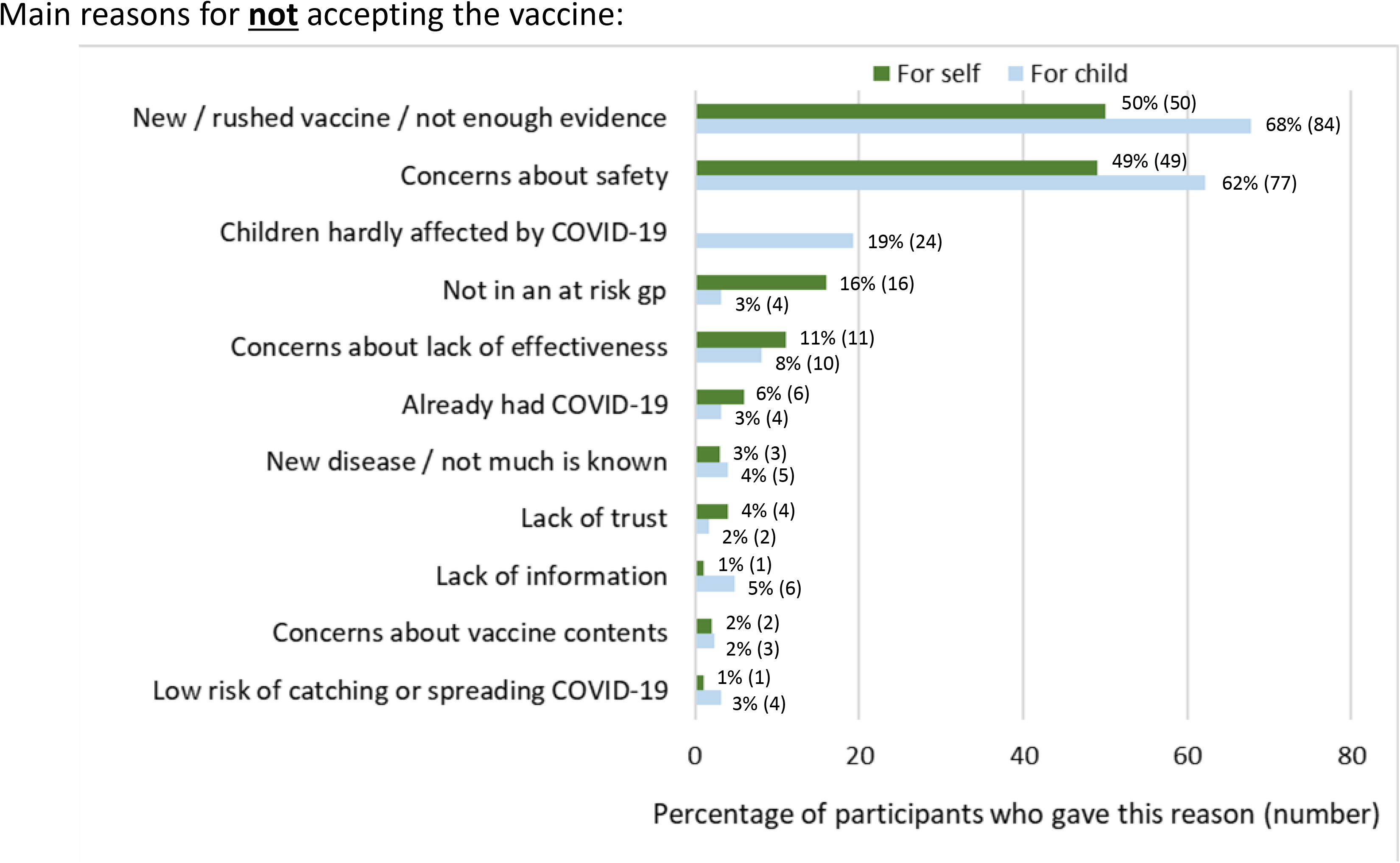

*Quantitative findings - Factors associated with COVID-19 vaccine rejection for self* A forward stepwise logistic regression analysis was performed with a dichotomised version of the self-vaccination against COVID-19 variable as the discrete variable. *Age, household income, ethnicity, location* and *employment were* included as predictive variables. The final model included three predictor variables *(household income, employment* and *ethnicity)* and significantly predicted ‘Likely to reject’ (omnibus chi-square = 50.225, df = 7, *p <.001). Age and location did not significantly predict ‘Likely to reject’, as such they are excl*uded from the model. The Hosmer-Lemeshow test demonstrates that the model adequately fits the data chi-square = 4.239, df = 7, *p* =.752. Table 4 gives coefficients and the Wald statistic, odds ratio and associated degrees of freedom for each of the predictor variables.

Participants in the lower household income bracket (<£35,000) were almost twice (OR: 2.08, 95%CI: 1.31 - 3.3) as likely to reject a COVID-19 vaccine than participants with a medium household income (£35,000-£84,999). Those in the highest income bracket (>£85,000) were almost three times (OR: 0.35, 95%CI: 0.17 - 0.73) as likely to accept the vaccine as middle-income bracket participants (£35,000-£84,999).

Participants that self-reported as Black, Asian, Chinese, Mixed or Other ethnicity were 2.7 times (95%CI: 1.27 - 5.87) more likely to reject a COVID-19 vaccine than White British, White Irish and White Other participants. There was also some indication that those that identify their employment as *homemaker* were more likely to reject the vaccination than those in full time employment or on parental leave from full time employment.

**Table 4.**

| Variable | Univariable analysis |  |  | Multivariable analysis |  |  |
| --- | --- | --- | --- | --- | --- | --- |
|  | Sig (p) | OR | 95% CI | Sig (p) | OR | 95% CI |
| **Age** | **.008** | - | - | **.559** | - | - |
| 18-23 | .017 | 3.575 | 1.257 – 10.165 |  |  |  |
| 24-30 | .004 | 1.860 | 1.216 – 2.844 |  |  |  |
| 31-36 † | - | - | - |  |  |  |
| 37-42 | .710 | .899 | .514 – 1.574 |  |  |  |
| 43+ | .439 | 1.634 | .471 – 5.669 |  |  |  |
| **Household income** | **<.001** | - | - | **<.001** | - | - |
| Low Income <£35,000 (n=242) | <.001 | 2.535 | 1.677 – 3.832 | .002 | 2.087 | 1.317 – 3.307 |
| Medium Income £35,000 - £84,999 (n=622)† | - | - | - | - | - | - |
| High Income >£85,000 (n=260) | <.006 | .363 | .177 - .744 | .005 | .352 | .170 – .729 |
| **Employment** | **<.001** | - | - | **.033** | - | - |
| Working full-time (n=628)† | - | - | - | - | - | - |
| Working part-time (n=373) | .802 | .943 | .596 – 1.491 | .229 | .739 | .452 – 1.209 |
| Homemaker (n=103) | <.001 | 3.025 | 1.784 – 5.130 | .038 | 1.888 | 1.035 – 3.445 |
| Student (n=10) | .006 | 5.673 | 1.656 – 19.436 | .311 | 2.076 | .506 – 8.524 |
| Unemployed (n=10) | .009 | 5.042 | 1.504 – 16.903 | .224 | 2.474 | .575 – 10.651 |
| **Ethnicity** | **.017** | - | - | **.010** | - | - |
| White (n=1,070)† | - | - | - | - | - | - |
| BAME (n=54) | .010 | 2.295 | 1.216 – 4.333 | .010 | 2.733 | 1.270 – 5.885 |

† Reference category

### Quantitative findings - Factors associated with COVID-19 vaccine rejection for child

A forward stepwise logistic regression analysis was performed with a dichotomised version of the child vaccination against COVID-19 variable as the discrete variable. *Age, household income, ethnicity, location, employment* and *number of children* were included as predictive variables. The final model included three predictor variables *(income, ethnicity* and *number of children)* and significantly predicted ‘Likely to reject’ (omnibus chi-square = 37.896, df = 6, *p* <.001). *Age, location* and *employment* did not significantly predict ‘Likely to reject’, as such they were excluded from the model. The Hosmer-Lemeshow test demonstrates that the model adequately fits the data chi-square = 1.502, df = 6, *p* =.958. Table 5 gives coefficients and the Wald statistic and associated degrees of freedom and probability values for each of the predictor variables.

Similarly, to accepting a novel covid-19 vaccination for themselves, participants that self-reported as Black, Asian, Chinese, Mixed or Other ethnicity were 2.74 times (95%CI: 1.35 - 5.57) more likely to reject a COVID-19 vaccine for their child than White British, White Irish and White Other participants. This finding was also found for income, with participants in the lower household income bracket (<£35,000) being 1.8 times (95%CI: 1.17 - 2.82) as likely to reject a COVID-19 vaccine for their child than participants with a medium household income (£35,000-£84,999). Participants with more than four children were also found to be around four times (OR 4.13; 95%CI: 1.873 - 9.104) more likely to reject the vaccination for their children than those with only one child, however, caution should be taken with this finding due to the small sample size.

**Table 5.**

| Variable | Univariable analysis |  |  | Multivariable analysis |  |  |
| --- | --- | --- | --- | --- | --- | --- |
|  | Sig (*p*) | OR | 95% CI | Sig (*p*) | OR | 95% CI |
| **Employment** | **<.001** |  |  | **.201** |  |  |
| Working full-time (n=628)† | - | - | - |  |  |  |
| Working part-time (n=373) | .923 | 1.021 | .666 – 1.568 |  |  |  |
| Homemaker (n=103) | <.001 | 2.561 | 1.511 – 4.340 |  |  |  |
| Student (n=10) | .074 | 3.377 | .891 – 12.805 |  |  |  |
| Unemployed (n=10) | <.001 | 8.684 | 2.829 – 26.660 |  |  |  |
| **Household income** | **<.001** |  |  | **.001** | - | - |
| Low Income <£35,000 (n=242) | <.001 | 2.291 | 1.521 – 3.451 | .007 | 1.826 | 1.178 – 2.830 |
| Medium Income £35,000 - £84,999 (n=622)† | - | - | - | - | - | - |
| High Income >£85,000 (n=260) | .094 | .614 | .347 – 1.087 | .079 | .596 | .334 – 1.062 |
| **Number of children** | **<.001** |  |  | **.004** | - | - |
| 1 (n=507)† | - | - | - | - | - | - |
| 2 (n=447) | .289 | 1.249 | .828 – 1.882 | .409 | 1.206 | .773 – 1.880 |
| 3 (n=133) | .014 | 1.955 | 1.146 – 3.335 | .083 | 1.677 | .935 – 3.009 |
| 4 or more (n=37) | <.001 | 4.250 | 2.044 – 8.836 | <.001 | 4.054 | 1.843 – 8.914 |
| **Ethnicity** | **.038** |  |  | **.009** | - | - |
| White (n=1,070)† | - | - | - | - | - | - |
| BAME (n=54) | .027 | 2.046 | 1.087 – 3.852 | .009 | 2.549 | 1.259 – 5.159 |

†Reference category

## Discussion

In this mixed methods study, the majority of parents and guardians reported they would definitely accept or were unsure but leaning towards accepting a COVID-19 vaccine for themselves (*Yes, definitely* 55.8%, n=699; *Unsure but leaning towards yes* 34.3%, n=429) and their child/children (*Yes, definitely* 48.2%, n=604; *Unsure but leaning towards yes* 40.9%, n=512). Less than 4% of parents and guardians reported that they would definitely not accept a COVID-19 vaccine for themselves or their children.

Attitudes towards a COVID-19 vaccine appear more positive in our study (in terms of the proportion of participants reporting they would definitely accept or were leaning towards accepting vaccination) than those reported in surveys conducted with adults in France, Germany, Italy, Portugal, the Netherlands and the US [12-14], and comparable to reports from Denmark and Australia [13, 15]. However, this could be due to differences in the questions asked; highlighting the need for cross-country surveys and consistency in the wording of questions. These surveys indicate that the difference in vaccination acceptance ranges greatly between countries, from around 62% in France to 80% in Denmark and the UK [13], and are reflective of trust in vaccines and health systems more broadly, and in governments. More positive attitudes towards COVID-19 vaccination in our study appear to reflect the high level of parent trust in vaccines in the UK, and trust in the NHS.

The main reasons, or ‘pros’ for vaccinating, were to protect the individual being vaccinated against COVID-19, followed by protecting others from the disease. Uncertainties around vaccine safety, effectiveness, and the benefit of vaccinating children were the most prevalent reasons given for COVID-19 vaccine refusal. Safety and effectiveness concerns were prompted by fears that the accelerated vaccine development process, considered politically motivated, could result in cutting corners. Comparably, previous research also suggests that people consider older vaccines safer than newer ones [16,17], and highlights the importance of transparency in communicating about the vaccine development process and of vaccination safety testing.

When a COVID-19 vaccine is launched, should any safety signals or safety concerns arise, it will be key for government and public health officials to reassure the public with transparency, action, accountability and timeliness in order to avoid any detrimental impact of the new vaccine on the current national immunisation programme. The Dengvaxia controversy in the Philippines highlights how safety concerns around a newly launched vaccine, Dengvaxia, can derail a successful national immunisation programme leading to drops in childhood vaccine uptake, and even spill over to other health care treatments [18].

Other surveys have identified that views on COVID-19 vaccine acceptance are influenced by vaccine efficacy and perceptions of disease risk. In a cross-sectional survey conducted in Indonesia, Harapan et al [19] found that 93.3% of participants would receive a COVID-19 vaccine that was 95% effective but only 67.0% of participants would accept a 50% effective vaccine. The authors also found that acceptance was higher amongst participants that considered themselves at greater risk of COVID-19 infection.

In our study, lower levels of vaccine acceptance for children were discussed by parents in terms of children being at lower-risk of COVID-19. Evidence suggests that children have lower susceptibility to COVID-19 [20], very few children develop severe COVID-19 (even if they are in a clinical risk group), and their role in the transmission of COVID-19 is unclear [21]. Similarly with influenza vaccination, parents and guardians in our study more often considered the perceived benefits to their child, rather than societal benefits, as a reason to vaccinate their children [22].

### Acting now to prevent inequalities in COVID-19 vaccine uptake

We found that participants of Black, Asian, Chinese, Mixed or Other ethnicity reported were more likely to reject a COVID-19 vaccine for themselves and their child/children compared to White participants (i.e. White British, White Irish or White Other). Comparably, lower levels of COVID-19 vaccine acceptability in Black African and lower income groups have been reported in US based surveys [14]. This is of concern given the evidence that Black, Asian and minority ethnic (BAME) groups and people living in the most deprived areas are at higher risk of acquiring COVID-19 infection and at increased risk of death from COVID-19 [21]. Inequalities in vaccination uptake amongst BAME communities [23-26] and lower income groups [26, 27] already exist for certain vaccines and a concerted effort must be made to understand factors affecting vaccine acceptance and prevent ethnic inequalities in uptake of a future COVID-19 vaccine. We were unable to explore ethnic differences in COVID-19 vaccine acceptance due to insufficient representation of ethnic minority groups in writing free-text responses and agreeing to participate in interviews.

### Strengths and limitations

The quality of the study was enhanced through the use of a multi-methods approach in which interview and open-text responses were used to develop insight into factors underpinning quantitative responses. Our study took place at the height of the COVID-19 pandemic in England and a survey repeated now that we are ‘past the peak’ of COVID-19 cases and deaths and lockdown has been eased may yield different responses. This has already been indicated in the second wave of a large European survey looking at COVID-19 vaccine attitudes [28]. This highlights the need for longitudinal studies to measure the acceptability of a COVID-19 vaccine at different intervals.

Our recruitment strategy, using social media, achieved a high number of responses (n=1252). Although geographically representative, our participants were not overly representative in terms of household income and ethnicity. This meant that it was not possible to explore differences in vaccination views by ethnicity and household income when looking at open-text responses and conducting interviews.

## Conclusion

The success of COVID-19 vaccination programmes will rely heavily on public willingness to accept the vaccine. The main concerns raised by parents in our study were around the safety and effectiveness of a ‘rushed’ and new COVID-19 vaccine. To alleviate these concerns, there needs to be clear communication and transparency as to how COVID-19 vaccines are developed and tested, and safety and efficacy information.

Opening up a conversation with members of the public at an early stage is key to understanding factors that may affect vaccine acceptability, and developing approaches to allay any concerns. This must happen in parallel with efforts to develop a COVID-19 vaccine. Importantly, efforts must be made to understand and address factors that may affect COVID-19 vaccine uptake in Black, Asian and minority ethnic groups and lower-income households who are disproportionately affected by COVID-19.

## Data Availability

Data will be made available upon request via LSHTM Data Compass - https://datacompass.lshtm.ac.uk/

## Ethical approval

Ethical approval was granted by the London School of Hygiene & Tropical Medicine Observational Research Ethics Committee (study reference: 21879).

## Acknowledgements

We would like to thank the baby and toddler group leads who helped share the online survey with potential participants. We are especially grateful for the time and contribution of all parents and guardians who took part in the study.

